# Trends in body mass index and blood pressure associations from 1989 to 2018: co-ordinated analysis of 145,399 participants

**DOI:** 10.1101/2020.11.06.20226951

**Authors:** David Bann, Shaun Scholes, Rebecca Hardy, Dara O’Neill

## Abstract

**Background:** High body mass index (BMI) is an important contributor to higher blood pressure (BP) levels and its deleterious consequences. However, the strength of this association may be context-specific and differ across time due to increases in medication use or secular changes in body composition. Thus, we utilised two independent data sources to investigate if associations between BMI and systolic BP (SBP) in Britain changed from 1989-2018.

**Methods:** We used 23 repeated cross-sectional datasets—the Health Survey for England (HSE) at ≥25 years (1994–2018; N=126,742); and three British birth cohorts (born 1946, 1958, and 1970) with outcomes available at 43-46 years (N=18,657). Anthropometry and BP were measured using standard protocols. We used linear and quantile regression to investigate cross-sectional associations between BMI and SBP.

**Results:** In HSE, associations were weaker in subsequent years, and this trend was most pronounced amongst older adults—after accounting for sex, treatment and education, the mean difference in SBP per 1 kg/m^2^ increase in BMI amongst adults ≥55 years was 0.75mmHg (95% CI: 0.60, 0.90) in 1994, 0.66mmHg (0.46, 0.85) in 2003, and 0.53mmHg (0.35, 0.71) in 2018. In cohorts, BMI and SBP associations were of similar magnitude in 1958 and 1970 cohorts and weaker in the 1946 cohort. Quantile regression analyses suggested that associations between BMI and SBP were present both below and above the hypertension threshold.

**Conclusion:** The consequences of BMI may differ across time and by age —associations between BMI and SBP appear to have weakened in recent decades, particularly in older ages. Thus, at older ages, this weakening strength of association may partly offset the public health impacts of increases in obesity prevalence. However, BMI remains positively associated with SBP in all adult age groups, highlighting the potential adverse consequences of the ongoing obesity epidemic.

## Introduction

High body mass index (BMI) is an important modifiable determinant of high blood pressure (BP), as evidenced by meta-analyses of observational studies^1^ and weight-loss reduction interventions.^2^ While providing precise estimates of association between BMI and BP, such studies have not elucidated whether the relationship between BMI and BP has changed across time. Evidence from two British birth cohorts born in 1946 and 1958 suggested an approximate doubling of the positive correlation between BMI and BP in midlife from 1989 to 2003.^3^ Other studies have reported increasing strength of association, yet used hypertension as the sole outcome^4 5^ (defined as elevated BP or use of BP lowering treatment) and thus may be simply reflecting increases in treatment use.^6^ In contrast, other studies have reported weakening of the association,^7 8^ or reported no clear systematic change.^6^

Understanding whether the associations between BMI and BP have changed across time is important to inform the future public health responses to obesity, the prevalence of which has increased markedly since the 1980s^9-11^ and may continue to increase. Indeed, projections suggest as many as 40-50% of the UK and US population may be obese by 2030.^12^ If the risks of rising BMI on raised BP levels are also increasing, then the public health impacts of BMI may be greater than anticipated given the importance of high BP for a range of cardiovascular^13^ and neurological outcomes.^14 15^

Multiple factors could explain why associations between BMI and BP may differ across time.^16^ Recent decades have seen improvements in hypertension awareness, detection, and treatment— notably via increased use of antihypertensive medication.^17^ Such interventions, along with behavioural changes triggered at least in part by hypertension diagnosis in primary care,^18^ may weaken such associations by lowering raised BP levels amongst those at highest cardiovascular risk, including those with higher BMI. However, hypertension treatment is only applicable to the relative minority (<25%^19^) of the population above specified hypertension treatment thresholds and even if initiated it may not fully normalise BP levels.^20 21^ Further, obtaining long-term changes to risk factors such as diet and physical activity is notoriously challenging. Other changes which may also impact on the strength of BMI and BP associations include secular changes to body composition—a high BMI may reflect more fat mass in more recent decades,^22 23^ and since high fat rather than high muscle mass is thought to impact on high BP^2^, associations may in turn strengthen across time.

We investigated whether BMI and BP associations systematically changed across time in Britain— from 1989 to 2018—employing data from two sources historically used separately: birth cohort studies and repeated cross-sectional studies. Each has complementary advantages. The birth cohorts have comparable BP measures obtained in midlife— a purportedly important period for cardiovascular risk^24-26^—and early life measures of potential confounding factors. Cross-sectional studies in turn collect data more regularly and generalise to a broader age span.

## Methods

We used two data sources 1) 23 repeated cross-sectional studies (Health Survey for England^27^), with anthropometrics and BP measured in samples of adults (annually from 1994 to 2018, except 1999 and 2004 where BP was measured only among minority ethnic groups); and 2) three British birth cohort studies of persons born in 1946 (1946c), 1958 (1958c) and 1970 (1970c), with anthropometrics and BP measured in midlife (in 1989, 2003, and 2016; see cohort profiles^28-30^). For each, participants gave verbal and/or written consent to be interviewed, visited by a nurse, and to have weight, height, and BP measurements taken during a home visit via standardised protocols. BMI was calculated as weight (kg) divided by the square of height (m^2^). The protocols for BP measurement have been described elsewhere.^31^ Briefly, each study used standardised protocols after 5 minute rest periods. Omron devices were used in 1958c (705CP) and 1970c (HEM-907), and in HSE from 2003 onwards. 1946c used the Hawksley random zero sphygmomanometer and HSE before 2003 used the Dinamap 8100 monitor. The 1946c and pre-2003 HSE data were converted to Omron measures using previously published regression equations based on calibration studies.^32 33^ Research ethics approval was obtained from relevant committees. Details of these studies and the measurement protocols are available elsewhere.^31^

The analytical sample size was 145,399—those with outcomes at age 43-46 years (cohorts; N=18,657) or 25 years and over (HSE; N=136,942 with BP data; N=126,742 with both BMI and BP data). The primary outcome for our study was the continuous level of systolic BP (SBP), chosen a priori given its strong link with cardiovascular disease and mortality;^13^ diastolic BP (DBP) was used as a secondary outcome.

### Analytical strategy

Our main analyses accounted for reported use of BP lowering treatment since changes in treatment use could potentially confound time-trend differences in BMI and BP associations.^6^ We accounted for the expected average effects of antihypertensive medication use on BP by adding a constant of 10mmHg (SBP) and 5mmHg (DBP) to the observed BP values amongst those on treatment. This approximates “underlying” BP (i.e. the BP individuals would have if they were not on treatment);^34^ this (constant addition) method has been found to reduce bias in the estimated effect of key determinants on continuous levels of BP due to the effects of antihypertensive medication use.^20 21^

The birth cohort and HSE datasets were analysed separately. Data from each source was analysed overall (i.e., pooled across cohorts / survey years) as well as separately by birth cohort / survey year. We first plotted histograms of BMI and BP, and then used linear regression to estimate mean difference in SBP per unit increase in BMI. Analyses were first adjusted for sex and then education attainment—a potential confounder (common cause of both BMI and BP levels^31 35^), included as a categorical term comprising four groups: degree/higher (reference), A levels/diploma, O Levels/GCSEs/vocational equivalent, or none.^36^ A formal test of change across time in the BMI and BP association was examined in the pooled HSE dataset by fitting interaction terms (a negative sign indicated a weakening association in subsequent years). Trend analysis was conducted separately in two time-periods (1994-2002 and 2003-2018) to examine the influence of change in BP device and introduction of non-response weights. In cohort studies, in which lifetime socioeconomic position (SEP) data were available, additional adjustment was made for social class at birth, maternal education in childhood, and own social class at 43-46y. HSE models additionally adjusted for age. Given body composition^37^ and treatment use differences by age,^17^ the BMI and SBP association may differ by age—as such, we conducted additional analyses in HSE in younger (25-54 years; N=73,750) and older age groups (55+ years; N=52,992). HSE analyses were also restricted to 40-49 year olds (N=25,941) to compare with cohort data. We additionally conducted analyses separately in men and women to investigate if time-trends in BMI and SBP associations differed by sex.

In addition to linear regression, quantile regression^38^ on data pooled across cohorts / survey years (HSE) was used to estimate whether the observed mean difference in SBP per unit increase in BMI were driven by differences at the upper or lower part of the SBP distribution; such analyses help inform the possible impact of treatment use on BMI and BP associations. Estimates were obtained and plotted at the 5^th^, 10^th^, 25^th^, 50^th^ (median), 75^th^, 90^th^, and 95^th^ quantiles. To facilitate interpretation we plotted the quantiles corresponding to the 140mmHg SBP threshold for initiating BP lowering treatment.^18^

To address data missingness, multiple imputation by chained equations was completed, with 20 imputations performed for the cohort pooled data and 20 each for the sex-stratified cohort datasets (including all SEP indicators). For the HSE data, weighting was used in lieu of multiple imputation. Starting from 2003, weights have been created to minimise bias from non-response (including non-participation in the nurse visit among those interviewed at the first stage); the relevant weights for analysing BP data were therefore used from 2003 onwards.

## Results

In later (2016/18) compared with earlier years (1989-2004), mean BMI and obesity prevalence (BMI ≥30kg/m^2^) were higher (Table 1), and the distributions of BMI were wider and more right-skewed (Supplementary Figure 1). In HSE but not cohort datasets, mean SBP and hypertension prevalence (140/90mmHg and/or on BP-lowering medication) were lower in later years—the distributions of SBP were more normally distributed than BMI in each year (Supplementary Figure 1). The use of BP-lowering medication was more frequent in later years in both data sources; in contrast, DBP levels were lower in both later cohorts and HSE survey years (Table 1).

**Table 1.**
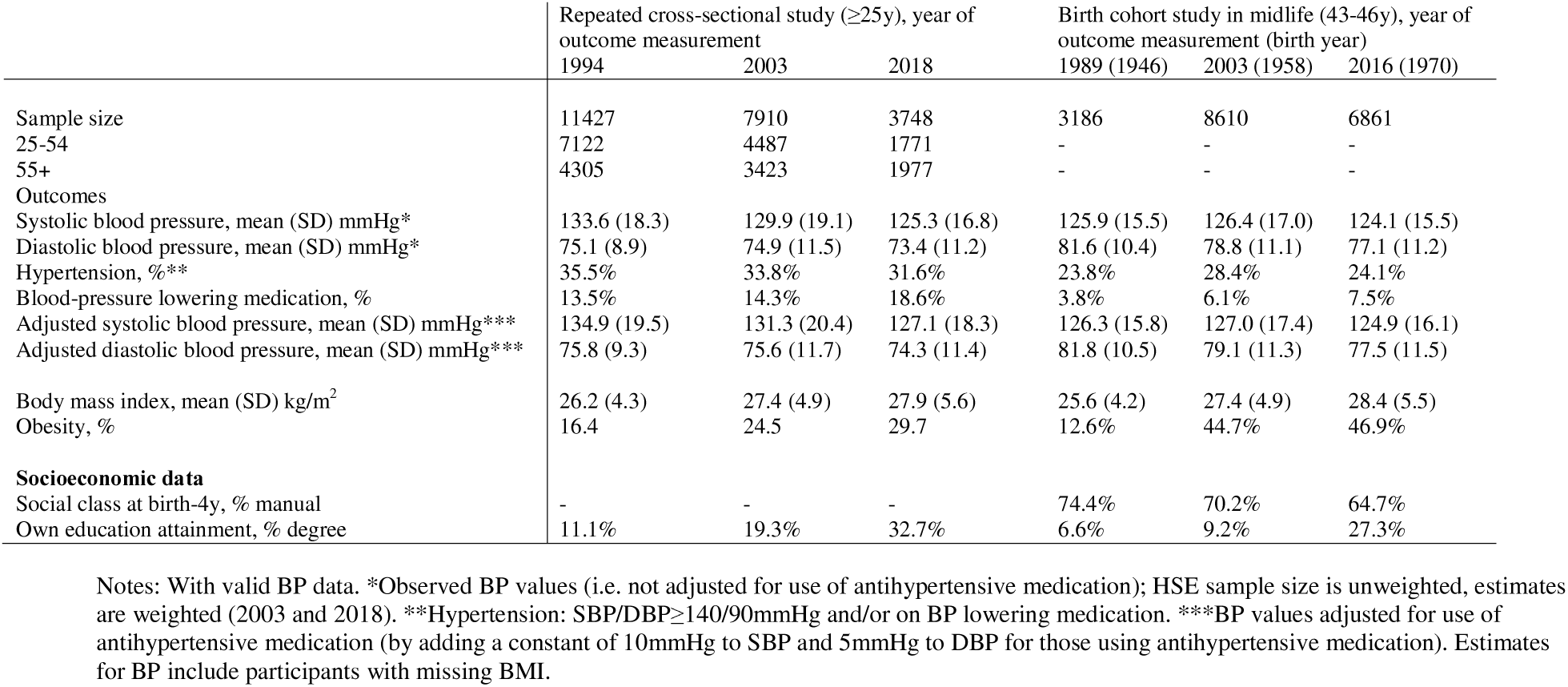
Participant characteristics: data from 23 repeated cross-sectional English studies and three British birth cohort studies.

| | Repeated cross-sectional study ($\geq 25$y), year of outcome measurement | | | Birth cohort study in midlife (43-46y), year of outcome measurement (birth year) | | |
| --- | --- | --- | --- | --- | --- | --- |
|  | 1994 | 2003 | 2018 | 1989 (1946) | 2003 (1958) | 2016 (1970) |
| Sample size | 11427 | 7910 | 3748 | 3186 | 8610 | 6861 |
| 25-54 | 7122 | 4487 | 1771 | - | - | - |
| 55+ | 4305 | 3423 | 1977 | - | - | - |
| Outcomes |  |  |  |  |  |  |
| Systolic blood pressure, mean (SD) mmHg* | 133.6 (18.3) | 129.9 (19.1) | 125.3 (16.8) | 125.9 (15.5) | 126.4 (17.0) | 124.1 (15.5) |
| Diastolic blood pressure, mean (SD) mmHg* | 75.1 (8.9) | 74.9 (11.5) | 73.4 (11.2) | 81.6 (10.4) | 78.8 (11.1) | 77.1 (11.2) |
| Hypertension, %** | 35.5% | 33.8% | 31.6% | 23.8% | 28.4% | 24.1% |
| Blood-pressure lowering medication, % | 13.5% | 14.3% | 18.6% | 3.8% | 6.1% | 7.5% |
| Adjusted systolic blood pressure, mean (SD) mmHg*** | 134.9 (19.5) | 131.3 (20.4) | 127.1 (18.3) | 126.3 (15.8) | 127.0 (17.4) | 124.9 (16.1) |
| Adjusted diastolic blood pressure, mean (SD) mmHg*** | 75.8 (9.3) | 75.6 (11.7) | 74.3 (11.4) | 81.8 (10.5) | 79.1 (11.3) | 77.5 (11.5) |
| Body mass index, mean (SD) kg/m<sup>2</sup> | 26.2 (4.3) | 27.4 (4.9) | 27.9 (5.6) | 25.6 (4.2) | 27.4 (4.9) | 28.4 (5.5) |
| Obesity, % | 16.4 | 24.5 | 29.7 | 12.6% | 44.7% | 46.9% |
| **Socioeconomic data** |  |  |  |  |  |  |
| Social class at birth-4y, % manual | - | - | - | 74.4% | 70.2% | 64.7% |
| Own education attainment, % degree | 11.1% | 19.3% | 32.7% | 6.6% | 9.2% | 27.3% |

Notes: With valid BP data. \*Observed BP values (i.e. not adjusted for use of antihypertensive medication); HSE sample size is unweighted, estimates are weighted (2003 and 2018). \*\*Hypertension: SBP/DBP ≥ 140/90mmHg and/or on BP lowering medication. \*\*\*BP values adjusted for use of antihypertensive medication (by adding a constant of 10mmHg to SBP and 5mmHg to DBP for those using antihypertensive medication). Estimates for BP include participants with missing BMI.

### Associations between BMI and SBP

BMI was positively associated with SBP in all HSE survey years and cohorts—the trends in this association (by survey year/cohort) were similar before and after accounting for treatment, education attainment, and (in cohorts) multiple SEP indicators across life (Supplementary Figure 2).

In HSE across all ages (≥25 years), BMI-SBP associations were weaker in subsequent years (Figure 1; BMI*year interaction term: β=-0.011 (95% CI: -0.014, -0.008)). In analyses stratified by age group the association between BMI and SBP was stronger in younger (25-54 years) compared with older (≥55 years) ages (Figure 2). However, a weakening of association across time was more pronounced among older adults—mean differences in SBP per 1 kg/m^2^ increase in BMI (after accounting for treatment and education) were 0.75mmHg (0.60, 0.90) in 1994, 0.66mmHg (0.46, 0.85) in 2003, and 0.53mmHg (0.35, 0.71) in 2018. Owing to the lower sample sizes in the older age group, confidence intervals were wider. Consistent with this interpretation, evidence for a weakening association in subsequent years was stronger in older (BMI*year interaction term: β =-0.010 (−0.015, -0.005)) compared with younger adults (β=-0.006 (−0.009, -0.003)), yet confidence intervals overlapped. Trend analysis conducted separately in 1994-2002 and 2003-2018 (when HSE changed BP measurement device and non-response weights were introduced) yielded similar findings (Supplementary Table 1). Across all time points there was considerable year-to-year variability in the magnitude of association such that comparisons restricted to two survey years may lead to misleading conclusions regarding long-term trends.

**Figure 1.**
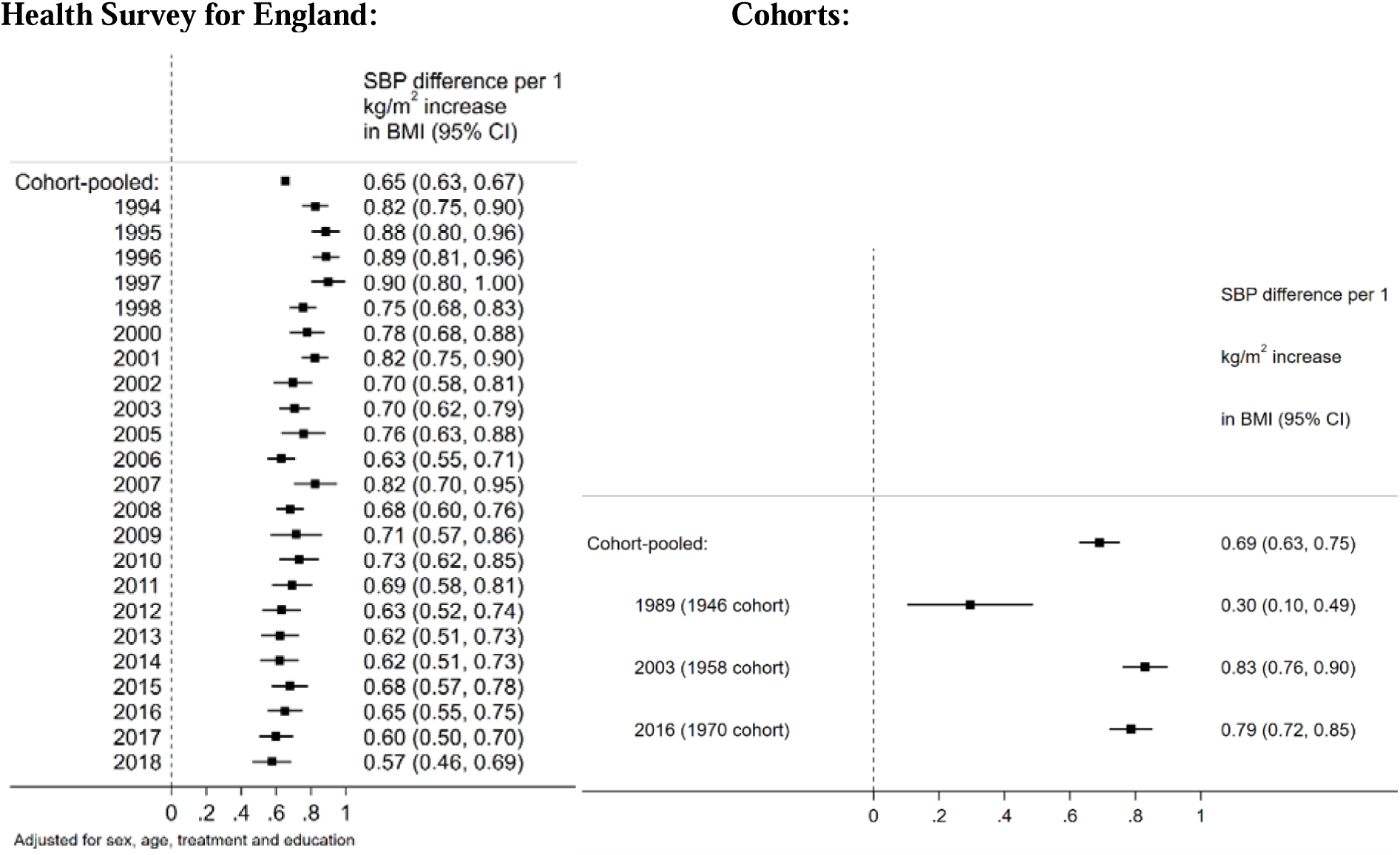
Associations between body mass index (kg/m^2^) and systolic blood pressure (mmHg) across adulthood (≥25 years, from repeated cross-sectional data, left panel), and midlife (43-46 years, from birth cohort data, right panel). Note: SBP values adjusted for use of antihypertensive medication (by adding a constant of 10mmHg for those using antihypertensive medication). Cohort models adjusted for mother’s education and cohort member’s own education, social class at birth, and social class in midlife.

**Figure 2.**
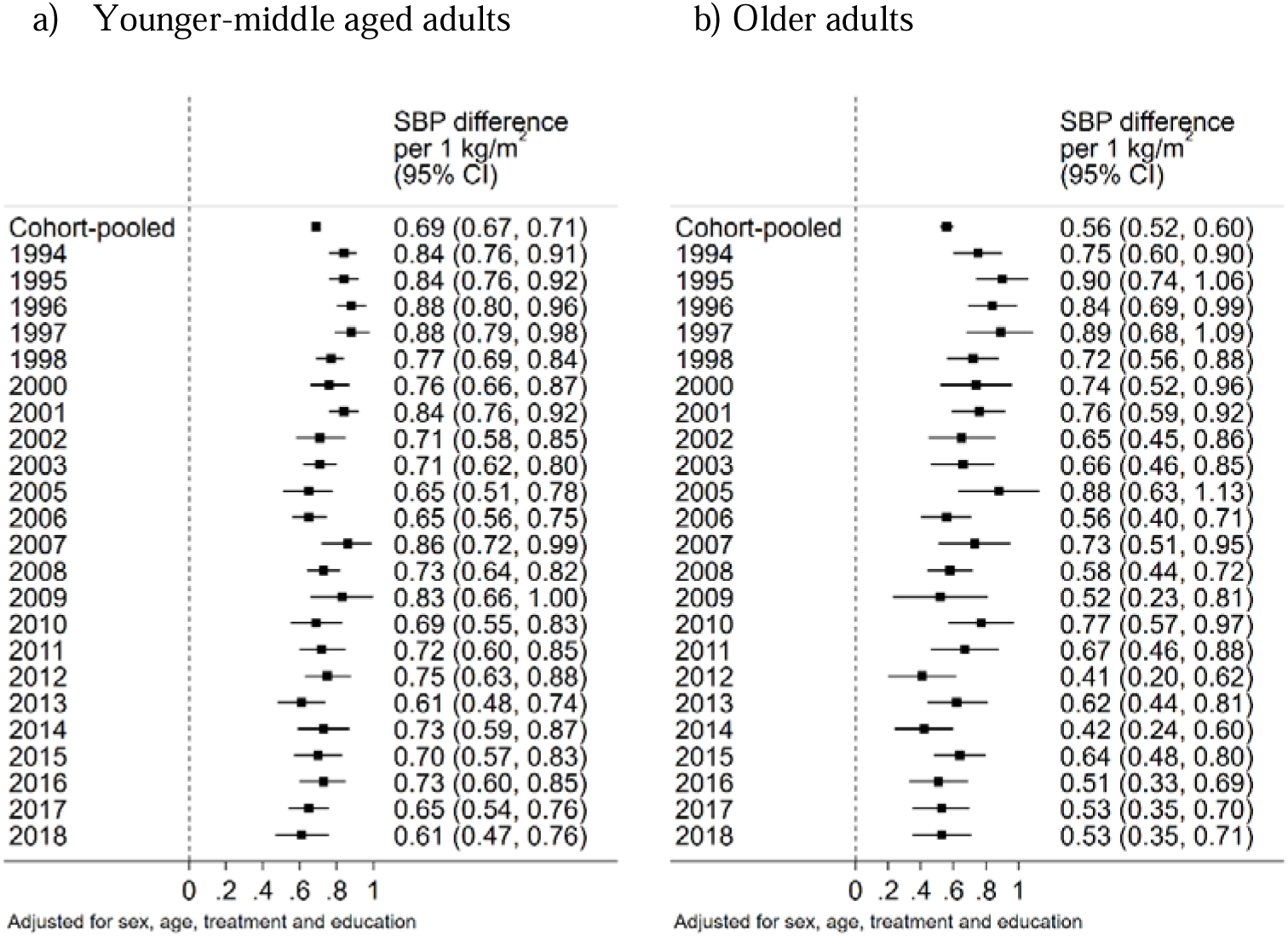
Associations between body mass index (kg/m^2^) and systolic blood pressure (mmHg) in 25-54 years (left panel) and 55+ years (right panel, from repeated cross-sectional data). Note: SBP values adjusted for use of antihypertensive medication (by adding a constant of 10mmHg for those using antihypertensive medication.

In cohorts (SBP measures at 43-46y only), the magnitude of association was substantially smaller in 1946c; magnitudes were highest in 1958c and slightly weaker in 1970c—the mean difference in SBP per 1 kg/m^2^ increase in BMI (after accounting for treatment and all SEP indicators) was 0.30mmHg (0.10, 0.49) in 1946c, 0.83mmHg (0.76, 0.90) in 1958c, and 0.79mmHg (0.72, 0.85) in 1970c (Figure Analyses using HSE found, in comparison with analyses in older adults (≥55 years), weaker evidence for systematic change among 40-49 year olds (Supplementary Figure 3; BMI*year interaction term, β=-0.002 (−0.007, 0.003)).

In HSE, the magnitude of associations between BMI and DBP were weaker prior to 2003, highest in 2003, and weaker in 2018 (Supplementary Figure 4). In cohort analyses, results using DBP yielded comparable results to those for SBP: associations between BMI and DBP were similarly weaker in 1946c, and higher in 1958c and 1970c (Supplementary Figure 4). Trends in the association between BMI and SBP were largely similar in men and women in HSE and cohort datasets (Supplementary Figure 5).

### Quantile regression analyses

Results of quantile regression analyses on data pooled across cohorts / survey years suggested that associations between BMI and SBP were present across the BP distribution—below and above the hypertension treatment threshold—yet were larger at upper SBP values (Figure 3 shows findings after accounting for treatment—findings were similar when not accounting for treatment). For example, in cohort-pooled analyses there was a 0.72mmHg (0.67-0.78) difference in SBP at the 50th quantile (median), per 1 kg/m^2^ increase in BMI, yet a 0.92mmHg (0.80-1.03) difference in SBP at the 90th quantile. Findings were similar in HSE (Figure 2).

**Figure 3.**
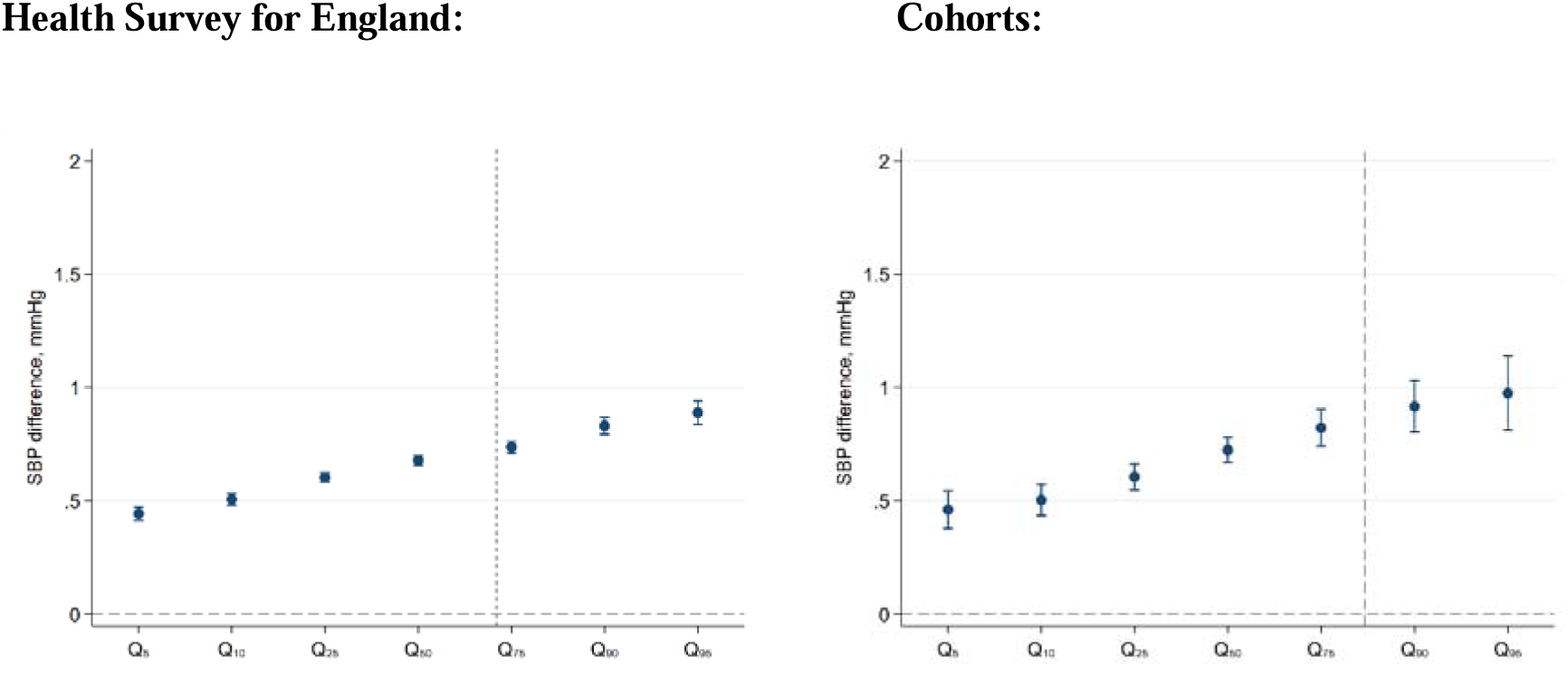
Associations between body mass index and systolic blood pressure quantiles (mmHg) across adulthood (≥25 years, from repeated cross-sectional data, left panel), and midlife (43-46 years, from birth cohort data, right panel). Note: SBP values adjusted for use of antihypertensive medication (by adding a constant of 10mmHg for those using antihypertensive medication; estimates are adjusted for sex and education attainment.

## Discussion

Using data from 23 cross-sectional datasets, we found that associations between BMI and SBP were weaker from 1994 to 2018; this trend was strongest in older (≥ 55 years) compared with younger (25-54 years) adults. Analysis of three birth cohorts with data in midlife (43-46 years) did not provide clear evidence of systematic trend in the association; in repeated cross-sectional data, there was weaker evidence for systematic change in the equivalent age group (40-49 years). However, BMI remained positively associated with SBP in all age groups, highlighting the future adverse consequences of the ongoing obesity epidemic.

The apparent discrepancy in findings between HSE and cohort results requires explanation. The fact that the increasing strength of the cross-sectional BMI-SBP associations previously reported (from 1946c to 1970c—ie, 1989 to 2000)^3^ were not found in 1970c, nor in the 23 HSE datasets used (1994-2018), suggests that this finding may be attributable to particular features of the 1946c rather than reflecting a long-term trend. This may include chance differences in findings due to the sampling— 1946c was sampled in one week of March 1946, and only married mothers were included.^39^ Unobserved differences in BMI and BP measurement protocols could have also contributed to such differences. More measurement error in calculating BMI would lead to a weakening of BMI-BP associations (due to regression dilution bias^40^), yet is an unlikely candidate for the cohort difference in association since similar protocols for measuring height and weight were used in each cohort. Notably, the BP devices used differed in each cohort. While the weaker BMI-BP association in 1946c was observed both before and after applying calibration equations to account for device differences (data available on request), it is possible that the calibration did not calibrate BP measures equivalently. Indeed, the mean difference in SBP per 1 kg/m^2^ increase in BMI at a later age in 1946c (53 years, when an Omron device was used) was substantially larger than at 43y (β (at 53 years) =1.09mmHg (0.91-1.27) after accounting for treatment, vs 0.34mmHg (0.14-0.53) at 43 years). Such discrepancies may also explain the finding that BMI-DBP associations were weaker in HSE prior to 2003, coinciding with a similar change in device. If the older BP devices are responsible for these weaker associations with BMI in the 1946c and in HSE 1994-2002, it suggests that differential error has occurred (e.g., systematic underestimation of BP among those with higher BMI, or overestimation amongst those with lower BMI). Datasets with BP measures obtained using multiple devices and measured anthropometrics are required to test this possibility—it has broader implications given the possibility that it may bias comparisons made within- (i.e., longitudinally) and between-cohorts.

Our study extends and potentially clarifies the equivocal existing literature which examined trends across time in cross-sectional BMI-BP associations.^3-8 41 42^ To our knowledge, the present study is the largest and most comprehensive investigation of this issue conducted thus far. We used two different studies with complementary features, accounted for treatment and multiple SEP indicators, and examined associations by age group. Two previous studies (in the US and Japan) reported an increasing strength of association.^4 5^ However, these and other studies^41^ used hypertension as an outcome (i.e., high BP and/or use of BP lowering treatment), and their findings may thus be solely explained by the secular trend of rising treatment use (increasing hypertension prevalence disproportionately among those at highest cardiovascular risk) rather than changes in the BMI and BP association per se.^6^

In studies that used continuous BMI and BP measures, weakening of association across time has also been observed in studies conducted in Germany (1998 to 2011),^8^ the Seychelles (1989 vs 2004),^7^ and the US (1993 to 2007 and 2005 to 2012, in some but not all model specifications).^6^ Another in contrast reported a strengthening relationship in Taiwan (1998 to 2006),^42^ yet this study was restricted to a ‘healthy sample’ aged 20-59yrs who were free of common chronic diseases and who were not on long-term medication (including antihypertensives). Finally, we extend the existing literature by utilising quantile regression analysis— findings of which suggest that the influence of BMI on SBP appears to be present across the SBP distribution (below and above the hypertension treatment threshold).

The magnitude of BMI and underlying BP associations is likely modified both by factors which could weaken the association across time (such as reduced salt intake amongst those with higher BMI) and those which strengthen it (such as a higher fat mass for a given BMI value in more recent decades). Our findings suggesting persistent associations from 2003 to 2018 amongst younger-middle aged adults implies that these differing processes may have offset each other, leading to similar magnitudes of association. In older adults, the relative balance of these factors seemingly led to a weakening of association across time. While we lack direct evidence for increases in fat mass percent in this study context, evidence from the US and elsewhere does suggest that it may have occurred along with the increasing obesity epidemic.^22 23^ Recent public health initiatives in the UK have included screening programmes for higher BP levels amongst those at highest CVD risk,^43 44^ leading to increases in hypertension diagnosis and treatment amongst those with high BMI.^45^ While treatment has increased, as observed in our data, it has seemingly had a modest effect on BMI and BP associations, particularly in younger age groups. This is evidenced by 1) our analysis accounting for treatment, and 2) quantile regression analyses suggesting that much of the association between BMI and SBP is present below the 140mmHg threshold for hypertension treatment. Changes in health behaviours may also have played a role in these processes. Salt intake has declined in the UK in recent decades,^46^ and this may have been particularly beneficial to those with higher BMI. Further weakening of the link between BMI and BP could feasibly be achieved by additional improvements to the risk factor profile amongst those with higher BMI—such as reduced salt and calorie intake, and increases in physical activity.

### Strengths and limitations

Strengths include the use of multiple birth cohort and cross-sectional datasets—each data source has independent sampling, and complementary strengths (e.g., annual data available across all ages in HSE, and more detailed SEP data in cohort studies with a larger sample size at midlife), enabling more robust inferences regarding the main findings than either data source in isolation.

While our analysis accounted for the potential confounding role of socioeconomic factors, we cannot exclude the possibility of other unmeasured confounding factors. Many plausible confounders—such as preceding ill health and behavioural factors which influence both BMI and BP—are likely to be partly but imperfectly captured by SEP given their social patterning. We also accounted for missing data, by using multiple imputation and analytical weights, yet similarly cannot exclude the possibility that unmeasured factors associated with non-response may have biased our findings. The introduction of analytic weights in the HSE series in 2003 means that comparison of trends in association before and after this year should be treated with caution. Finally, while the contribution of treatment use was estimated to be minimal, our approximation of its effect on BMI-BP associations is imperfect owing to a lack of data on treatment, dose, and adherence.^6^

### Conclusion

The consequences of BMI may differ by both time period and age group—associations between BMI and SBP appear to have weakened in recent decades, particularly at older ages. This trend is less evident at younger ages—assuming causal relations between BMI and SBP, given increases in obesity prevalence in this age group, the population impact of BMI on SBP levels may be greater now than in the past. At older ages, the weakening strength of association may offset the impact of increases in obesity prevalence at the population level. However, BMI remains positively associated with SBP in all age groups, highlighting the future adverse consequences of the ongoing obesity epidemic. Finally, our results highlight the utility of using multiple datasets to obtain robust inferences regarding trends in risk factor-outcome associations across time; and the potential limitations of drawing inferences on long-term trends when comparing only two time-points.

## Data Availability

Data are available upon request at the links provided

https://www.nshd.mrc.ac.uk/data/data-sharing/

https://www.data-archive.ac.uk

