## Supplemental file for "Trends in body mass index and blood pressure associations from 1989 to 2018: co-ordinated analysis of 145,399 participants"

#### Observed data distributions

i) Cohort studies

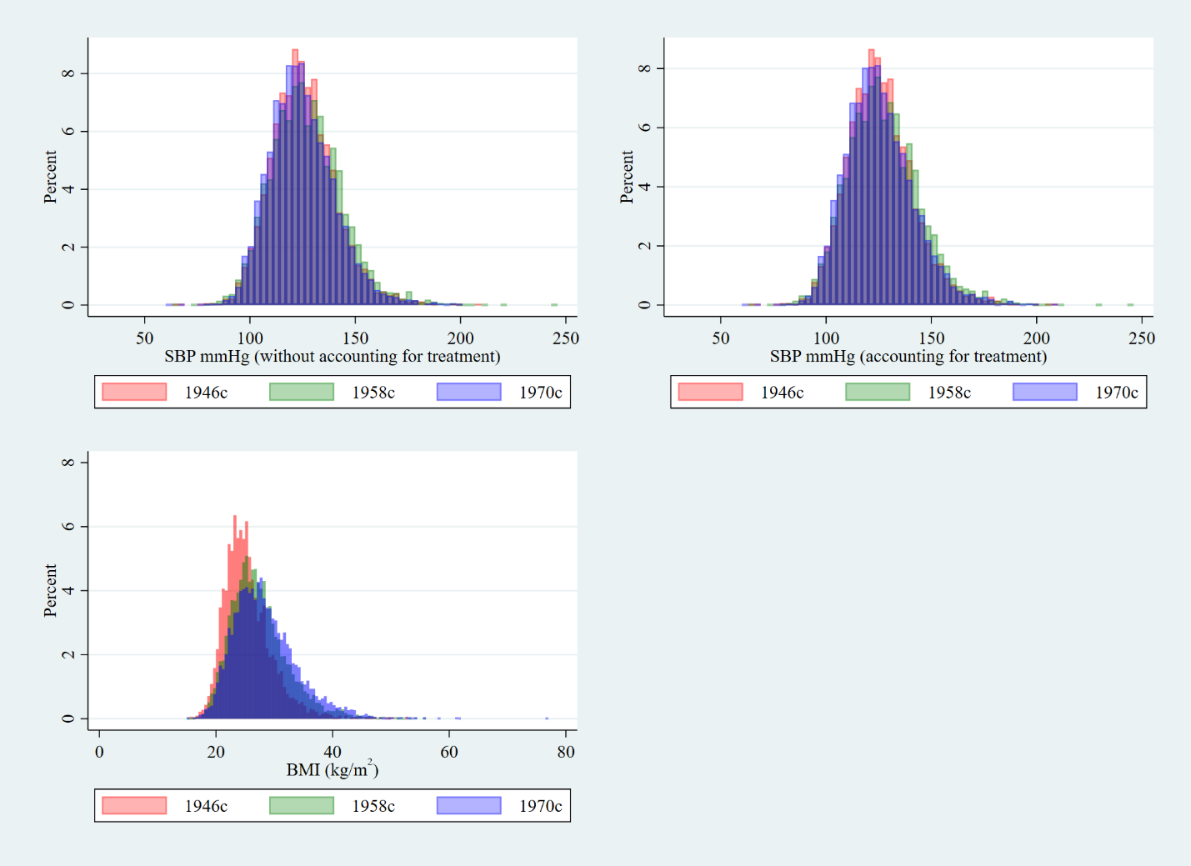

ii) Cross-sectional studies:

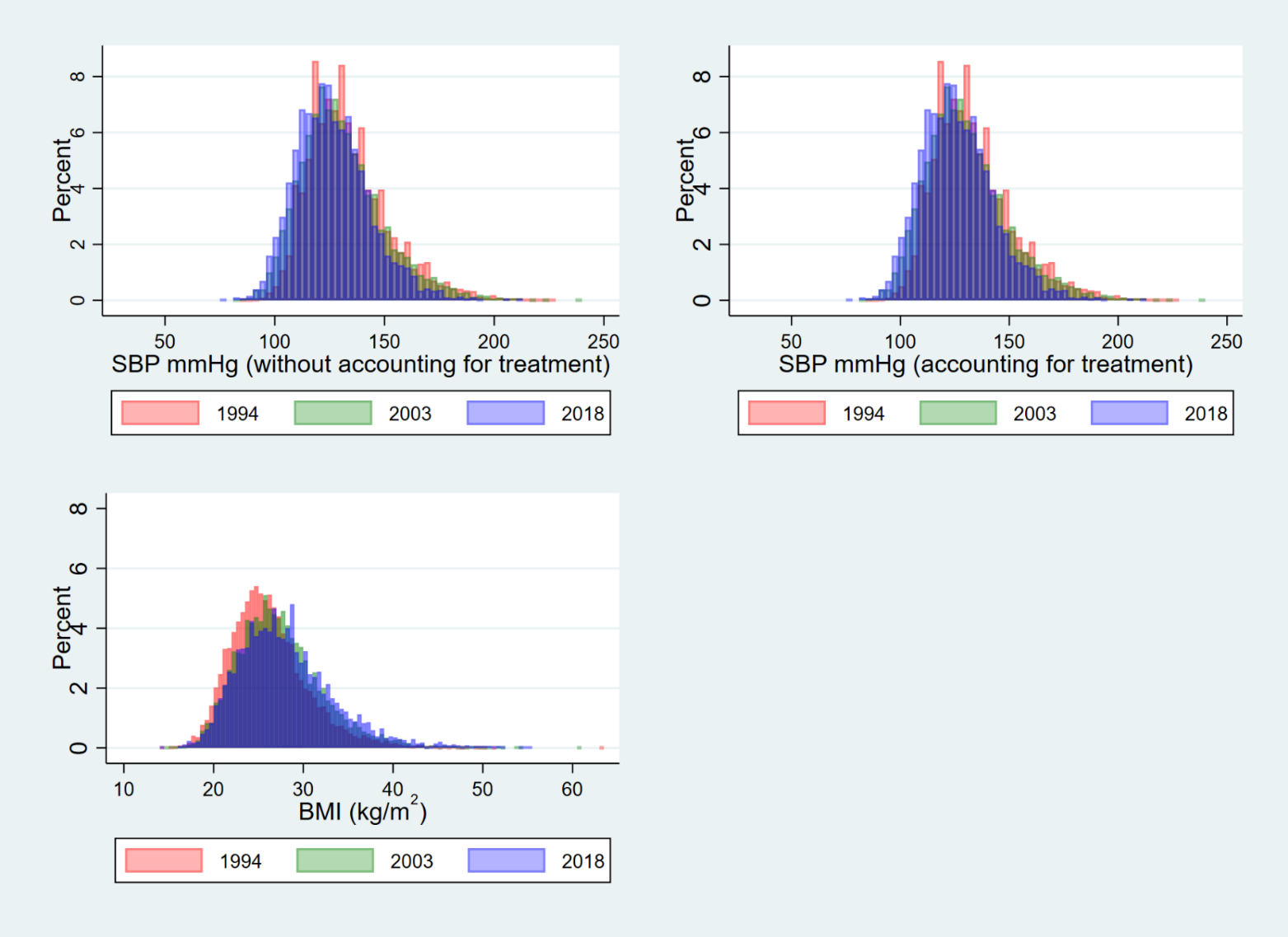

**Supplementary Figure 1. Histograms of observed SBP (mmHg, before and after accounting for hypertension treatment) and observed BMI in i) 1946, 1958 and 1970 birth cohorts and ii) repeated cross-sectional data in 1994, 2003, and 2018**

#### Analyses showing sex-adjusted associations before and after accounting for treatment and socioeconomic factors

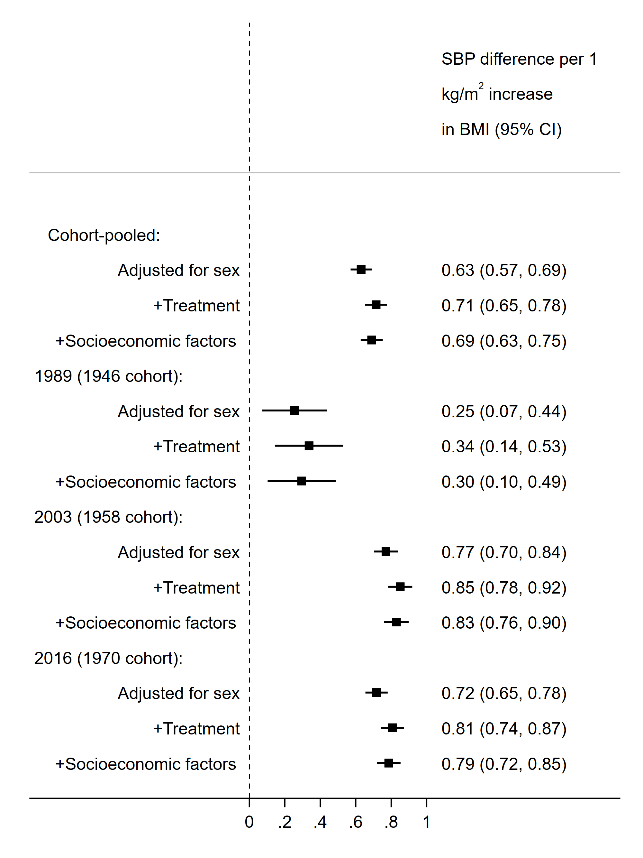

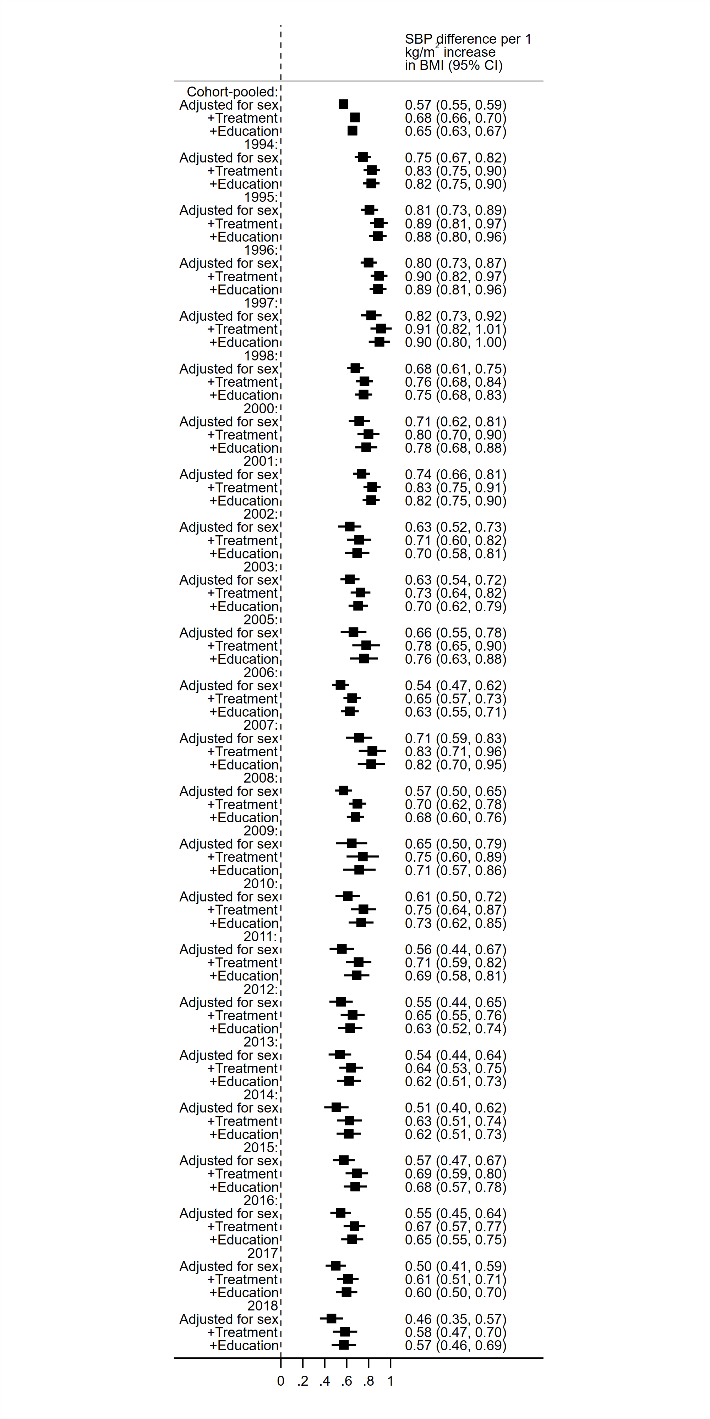

Supplementary Figure 2. Associations between body mass index (kg/m^2^) and systolic blood pressure (mmHg) in midlife (42-46 years, from birth cohort data, left panel) and across adulthood (≥25 years, from repeated cross-sectional data, right panel). Note: SBP accounted for treatment use by adding a constant of 10mmHg to those using antihypertensive medication. Socioeconomic factors comprise mother’s education and cohort member’s own education, social class at birth, and midlife social class.

#### Cross-sectional data restricted to 40-49 years age:

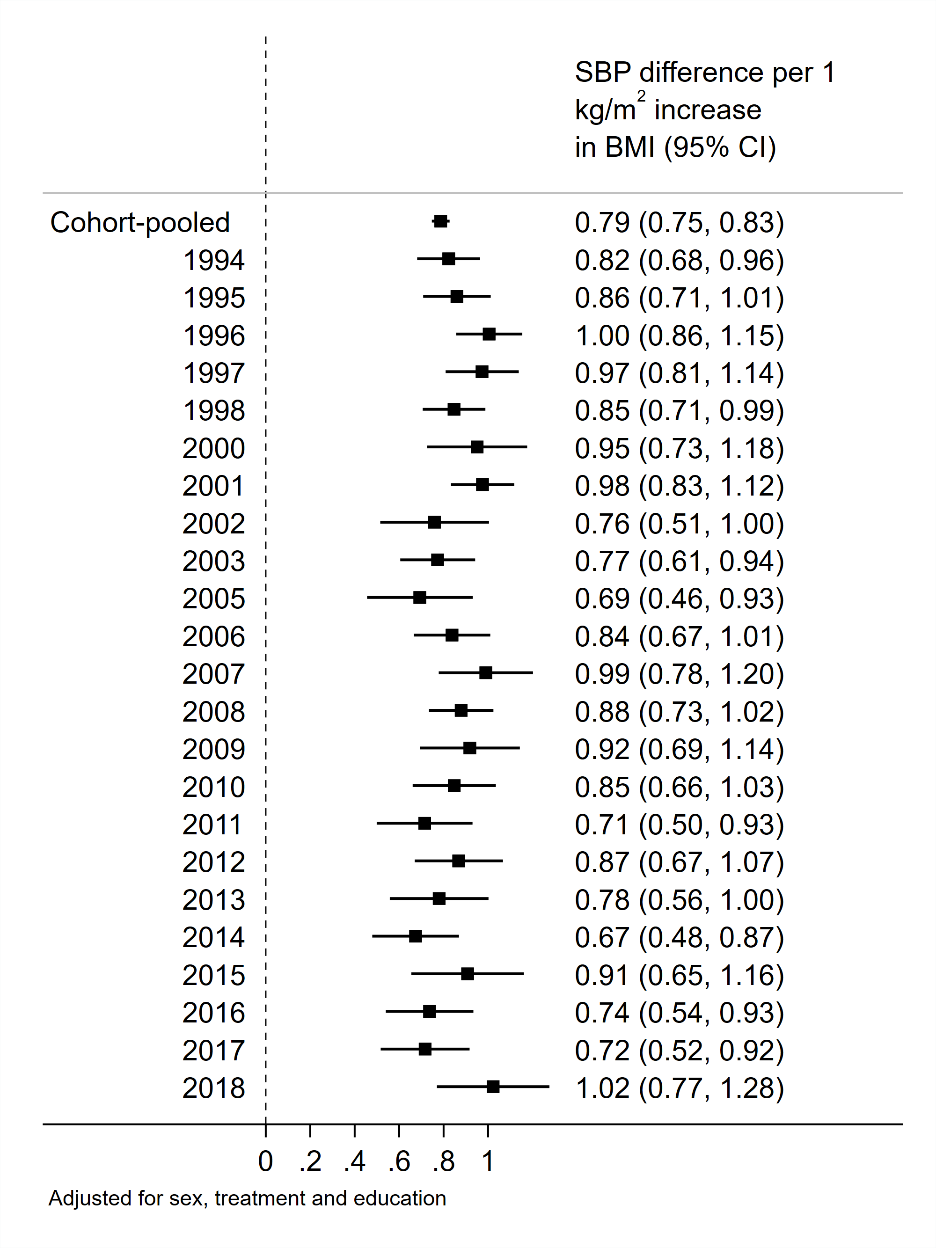

**Supplementary Figure 3. Associations between body mass index (kg/m^2^) and blood pressure (mmHg) in midlife (from repeated cross-sectional data).** Note: BP adjusted for treatment obtained by adding a constant of 10mmHg to SBP and 5mmHg DBP to those using antihypertensive medication.

Diastolic blood pressure

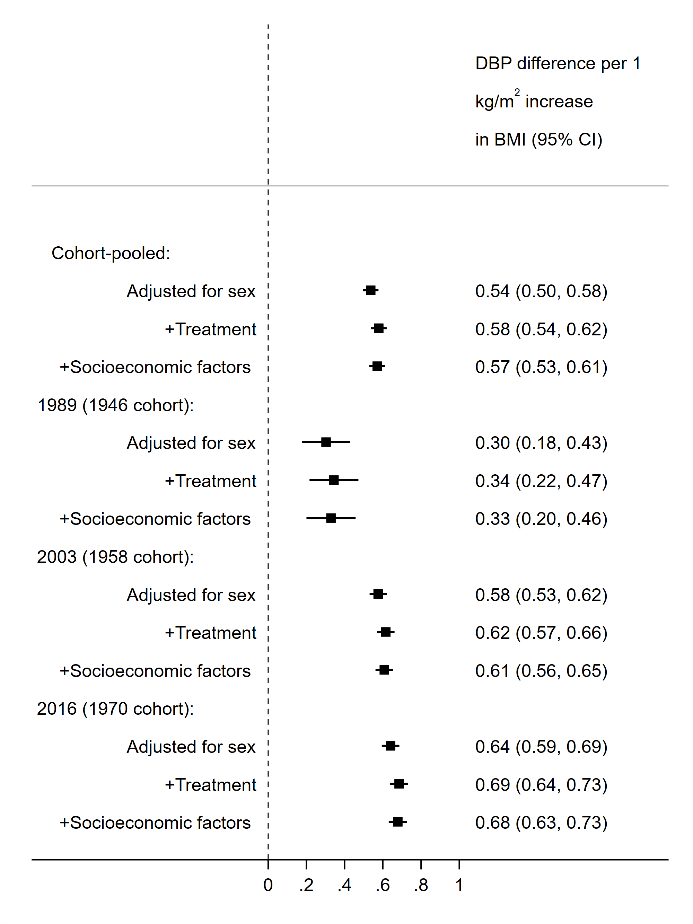

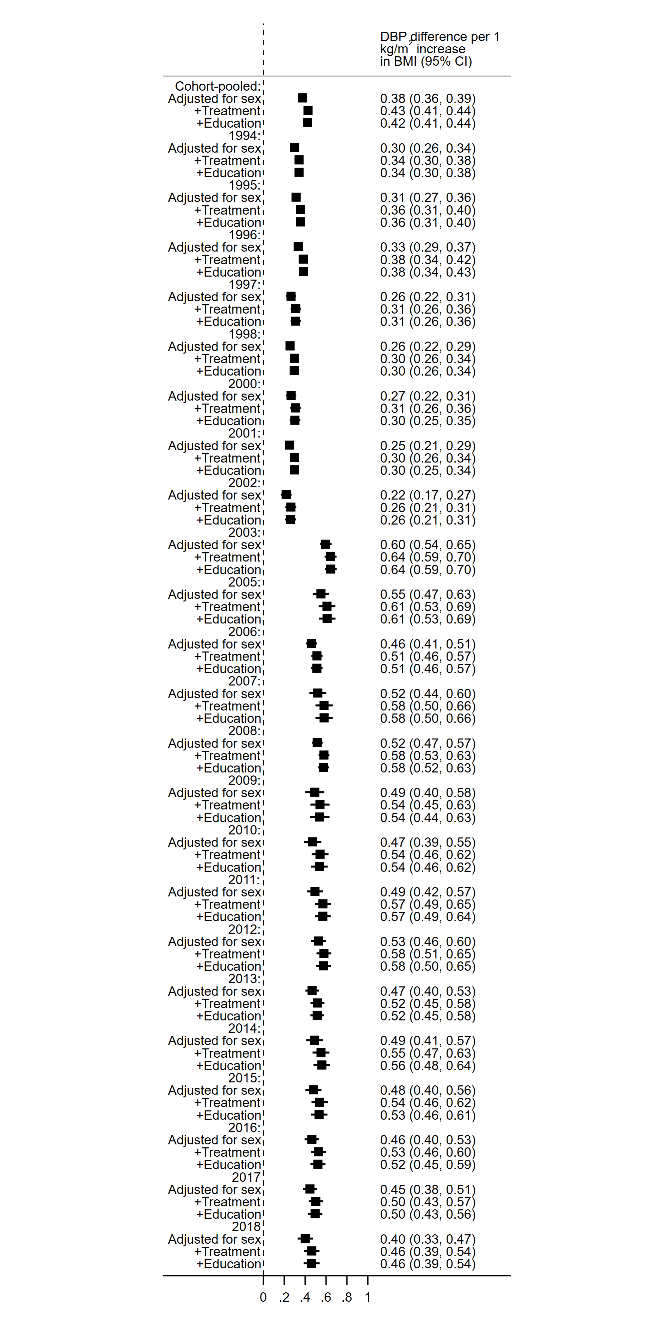

**Supplementary Figure 4. Associations between body mass index (kg/m^2^) and diastolic blood pressure (mmHg) in midlife (42-46 years, from birth cohort data, left panel) and across adulthood (≥25 years, from repeated cross-sectional data, right panel).** Note: DBP adjusted for treatment obtained by adding a constant of 5mmHg to those using antihypertensive medication. Socioeconomic factors comprise mother’s education and cohort member’s own education, social class at birth, and midlife social class.

#### Sex-stratified (Birth cohort data): Systolic blood pressure

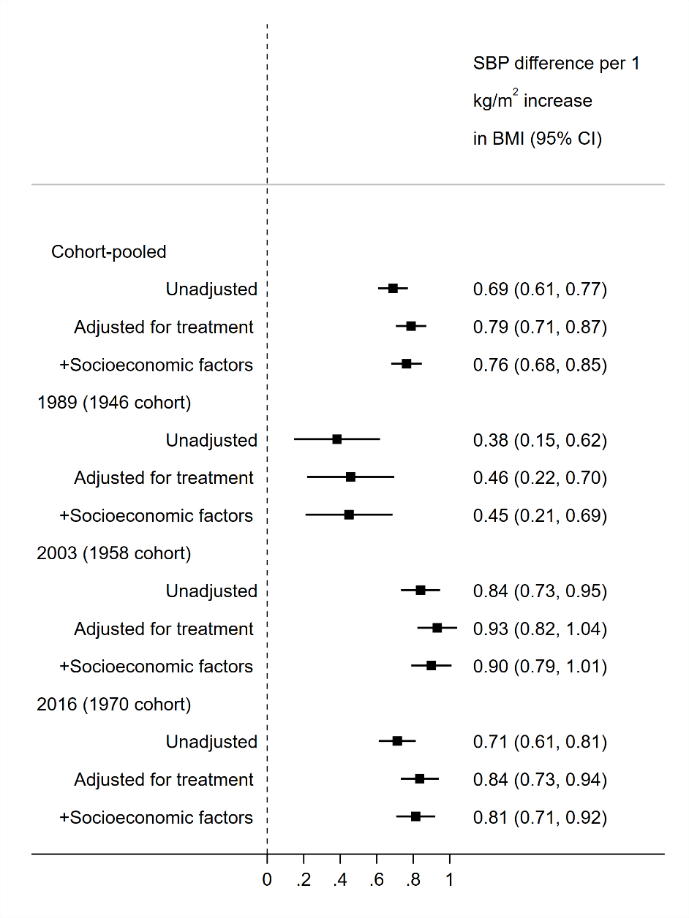

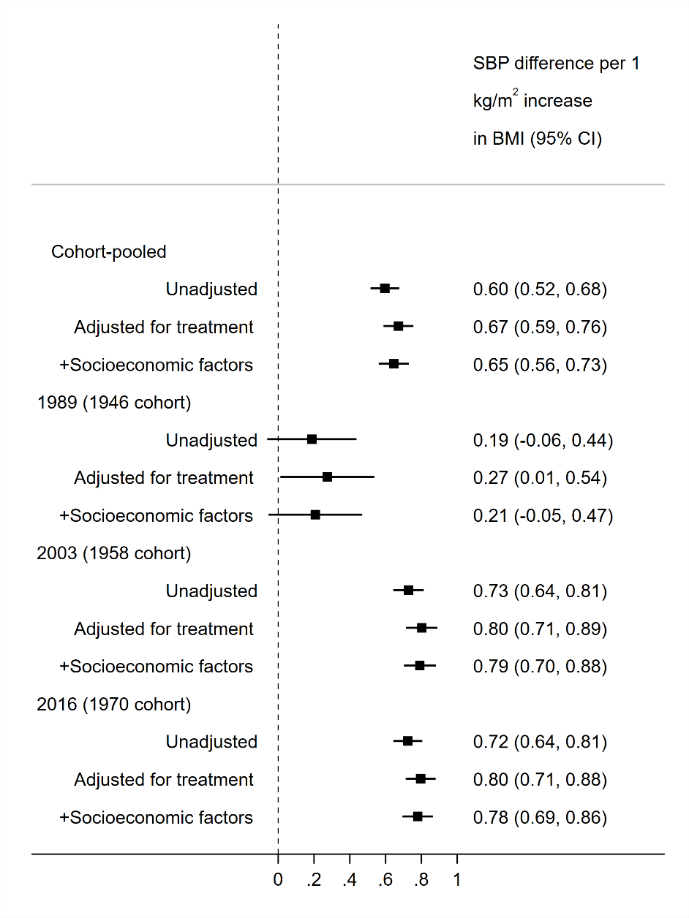

**Supplementary Figure 5. Associations between body mass index (kg/m^2^) and systolic blood pressure (mmHg) during midlife (42-46 years) in males (left panel) and females (right panel).** Note: BP adjusted for treatment obtained by adding a constant of 10mmHg to SBP to those using antihypertensive medication. Socioeconomic factors comprise mother’s education and cohort member’s own education, social class at birth, and midlife social class.

#### Sex-stratified (Repeated cross-sectional data, ≥25 years): Systolic blood pressure

…Supplementary Figure 5 continued.

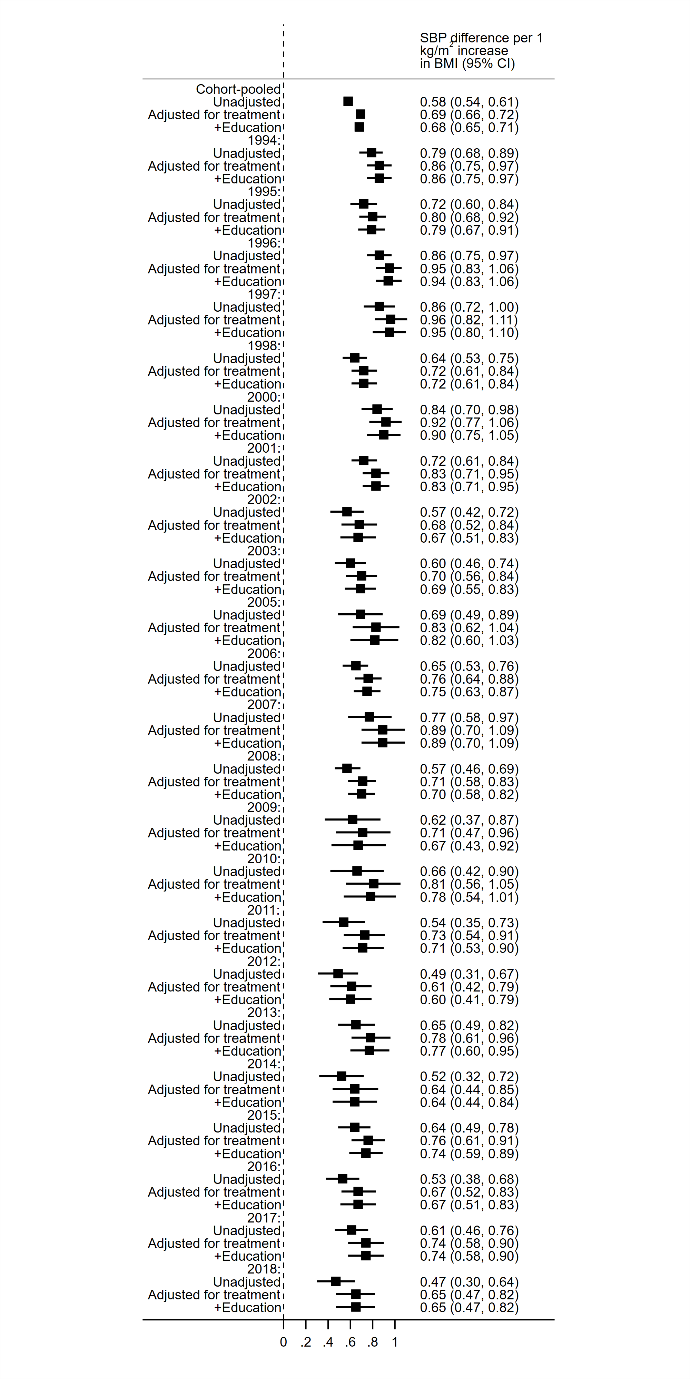

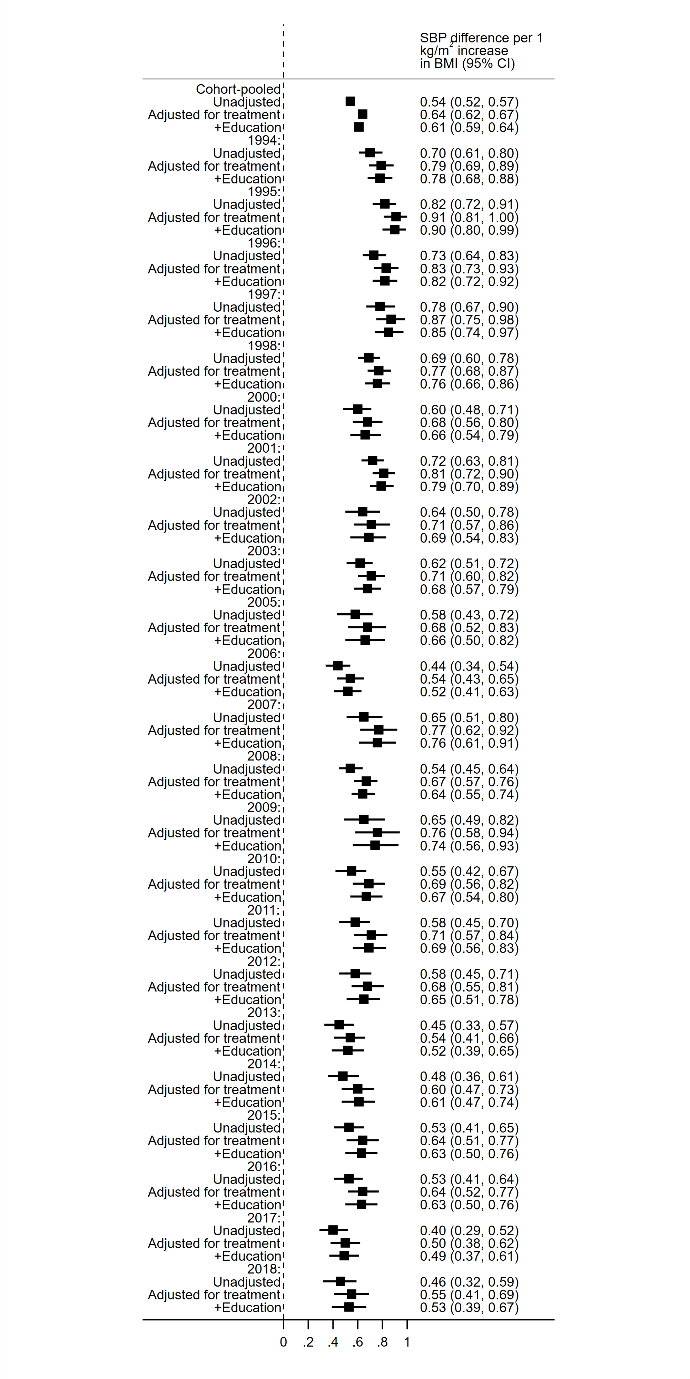

#### Sex-stratified (Birth cohort data): Diastolic blood pressure

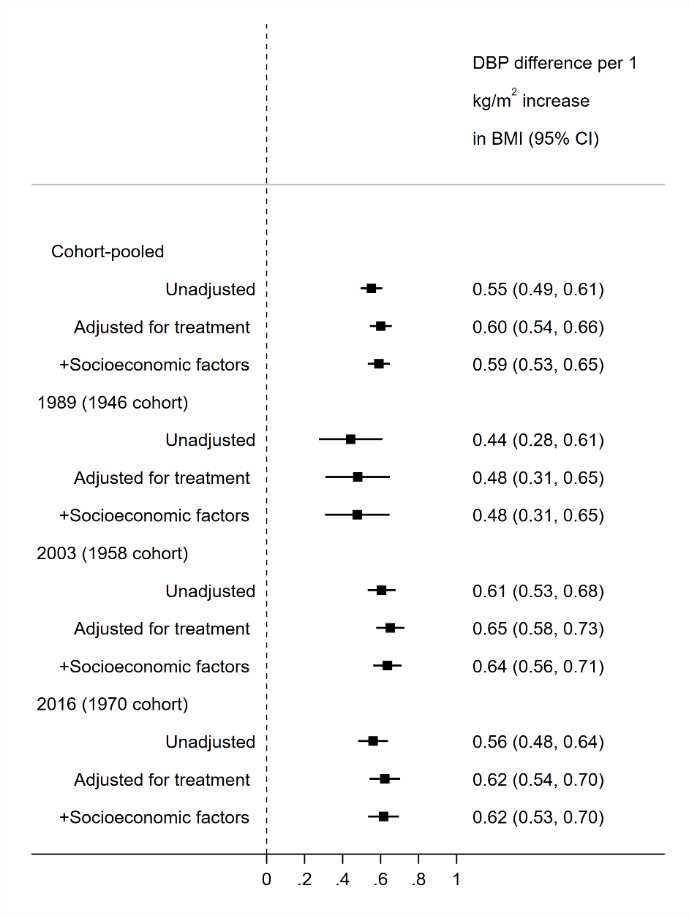

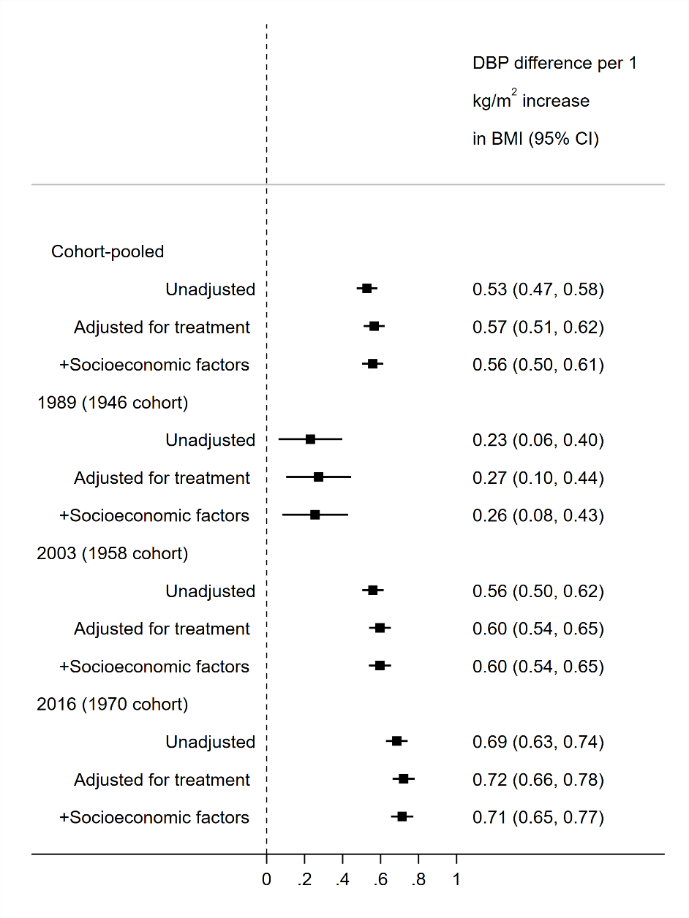

…Supplementary Figure 5 continued.

#### Sex-stratified (Repeated cross-sectional data, ≥25 years): Diastolic blood pressure

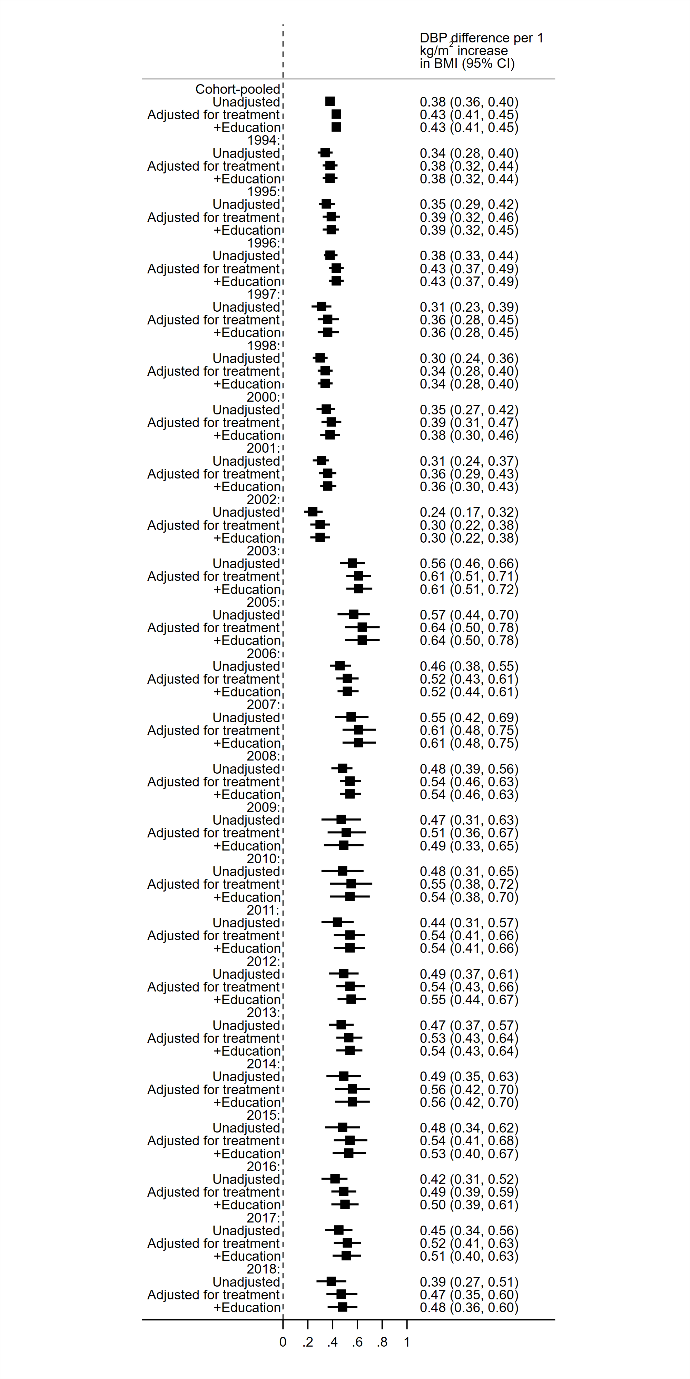

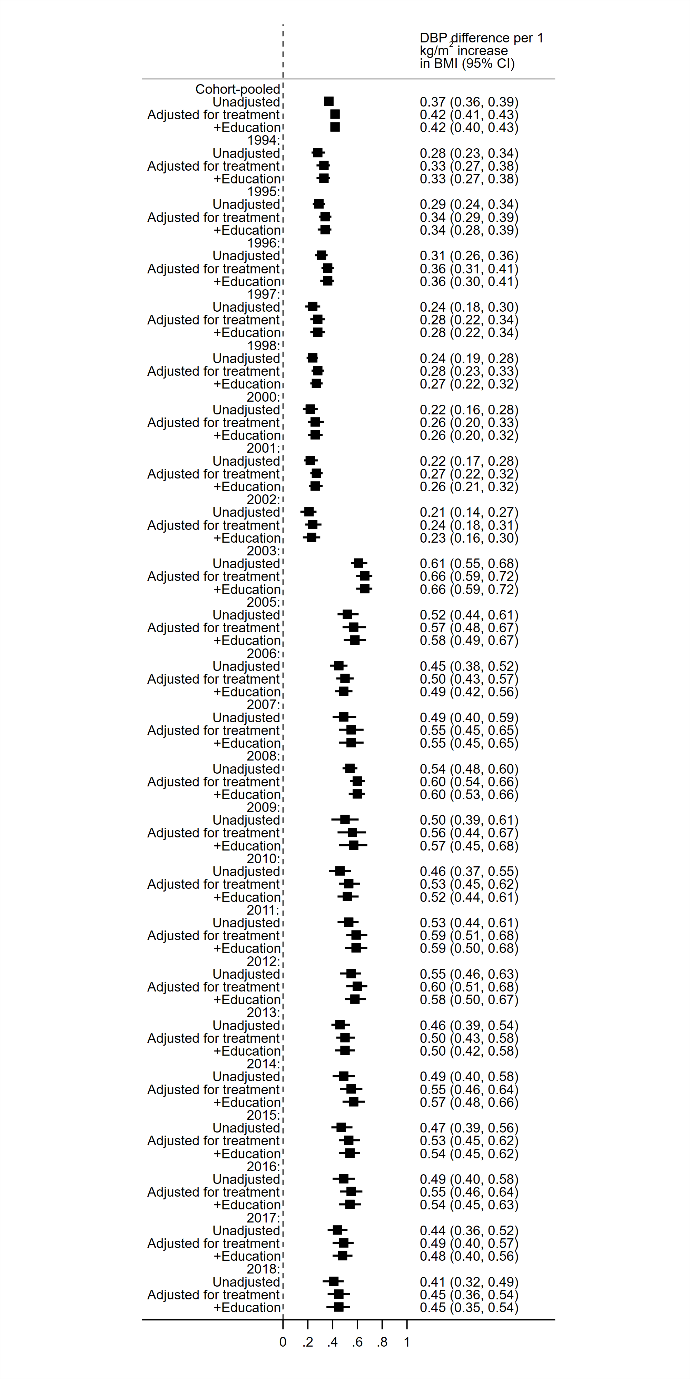

…Supplementary Figure 5 continued.

### Results of trend analyses—interaction terms for BMI*year interaction terms

**Supplementary Table 1.** Trends in BMI and SBP associations in the Health Survey for England, by age and year group**.**

| Slope for BMI*Year |  |  |  |  |  |
| --- | --- | --- | --- | --- | --- |
|  | Estimate | LL | UL | P-value | N |
| **25+:** |  |  |  |  |  |
| All years | -0.011 | -0.014 | -0.008 | <0.001 | 126,742 |
| Pre-2003 | -0.016 | -0.028 | -0.005 | 0.004 | 62,700 |
| 2003 & onwards | -0.009 | -0.015 | -0.004 | 0.001 | 64,042 |
| **25-54:** |  |  |  |  |  |
| All years | -0.006 | -0.009 | -0.003 | <0.001 | 73,750 |
| Pre-2003 | -0.010 | -0.021 | 0.002 | 0.106 | 39,267 |
| 2003 & onwards | -0.005 | -0.011 | 0.002 | 0.177 | 34,483 |
| **55+:** |  |  |  |  |  |
| All years | -0.010 | -0.015 | -0.005 | <0.001 | 52,992 |
| Pre-2003 | -0.013 | -0.036 | 0.010 | 0.27 | 23,433 |
| 2003 & onwards | -0.010 | -0.020 | 0.001 | 0.086 | 29,559 |

Notes: estimates obtained from separate linear regression models with SBP as the outcome, after accounting for treatment use and adjusting for education attainment.
